# Behavioural Outcomes of Preschool Children with Congenital Heart Disease and Controls

**DOI:** 10.64898/2025.12.04.25341634

**Authors:** Andrew TM Chew, Alexandra F. Bonthrone, Mirthe E. M. van der Meijden, Zeyuan Sun, Kuberan Pushparajah, John Simpson, A David Edwards, Serena J. Counsell, Chiara Nosarti

## Abstract

**Background:** Behavioural outcomes may be suboptimal in school-age children and adolescents with congenital heart disease (CHD), but little is known about outcomes of preschool children. We aimed to compare the behavioural outcomes of preschool children with CHD with controls, and investigate whether a cognitively stimulating home environment impacts their outcomes.

**Methods:** This is a cross-sectional case-control study of fifty-six preschool children (4-6 years) with CHD and 215 controls. Questionnaires were used to assess temperament (Child Behavior Questionnaire), autism traits (Social Communication Questionnaire), ADHD symptoms (ADHD-Rating Scale-IV), empathy (EmQue), and behavioural difficulties (Strengths and Difficulties Questionnaire). The Cognitively Stimulating Parenting Scale was used to assess home environment.

**Results:** Univariate robust regression analyses showed that children with CHD compared to controls had higher levels of hyperactivity-impulsivity (B=-0.467 p=0.023), inattention (B=-0.450, p=0.023) and peer relationship problems (B=-0.504, p=0.023), after controlling for age at assessment, gestational age at birth, sex and neighbourhood deprivation, with results surviving false discovery rate correction. Group significantly moderated the relationship between cognitively stimulating opportunities at home and the outcomes that differed between the groups: hyperactivity-impulsivity, inattention and peer problems. More cognitively stimulating opportunities at home were associated with more favourable behavioural outcomes in children with CHD (hyperactivity-impulsivity: B=-0.092, p<0.001; inattention: B=-0.088, p<0.001; peer problems: B=-0.124, p<0.001) but not in controls (hyperactivity-impulsivity: B=-0.005, p=0.727; inattention: B=-0.019, p=0.225; peer problems: B=-0.002, p=0.911).

**Conclusions:** Compared to controls, preschool children with CHD have more hyperactivity-impulsivity, inattention and peer relationship problems. These behavioural problems are mitigated by a cognitively stimulating home environment.

## Introduction

Congenital heart disease (CHD) remains the most common congenital anomaly affecting approximately 1% of livebirths worldwide (EUROCAT, 2025; Liu et al., 2019). Children with CHD are at increased risk of neurodevelopmental impairments and behavioural difficulties, with severity increasing with disease complexity (Davidson et al., 2015). As children enter preschool, they encounter environments that require greater independence and self-regulation hence the frequency and severity of behavioural difficulties may become more apparent (Green et al., 2023).

Previous studies have identified a wide range of suboptimal behavioural outcomes in school age children and adolescents with CHD, with a predominance of internalizing over externalizing symptoms (Abda et al., 2019; Bellinger et al., 2009; Latal et al., 2009; Majnemer et al., 2008), poorer emotional control, problems with self-esteem and body image, social interactions, and social cognition (Bar-Mor et al., 2000; Brudy et al., 2021; Chong et al., 2018; Pike et al., 2012; Salzer-Muhar et al., 2002). There is also increasing evidence that children with CHD are at greater risk of inattention or being diagnosed with attention deficit hyperactivity disorder (ADHD) (Czobor et al., 2021; DeMaso et al., 2014; Holland et al., 2017; Holst et al., 2020; Tang et al., 2024). Autism diagnosis or traits are also increased in school-age children, adolescents and young adults with CHD (Asschenfeldt et al., 2020; Bellinger et al., 2011; Neal et al., 2015; Razzaghi et al., 2015).

The behavioral outcomes of preschool children with CHD are not as well researched as those of school-age children and adolescents (Abda et al., 2019; Gu et al., 2023; Hasan et al., 2023). The preschool years are a time of rapid socioemotional development, marked by rapid changes in development, laying the foundation for future learning in school (Ansari, 2018). The studies assessing behavioral differences in preschool children with CHD show conflicting results, although these are difficult to compare directly due to methodological differences. Some studies have reported no behavioral differences in preschool children with CHD compared to population norms or controls (Brosig et al., 2007; DeMaso et al., 1990; Goldberg et al., 1997; Hövels-Gürich et al., 2001; Majnemer et al., 2006; Stene-Larsen et al., 2011; Utens et al., 2001), while a few have reported significant differences in emotional problems or autism traits (Bean Jaworski et al., 2017; Gaynor et al., 2009; Hülser et al., 2007) and ADHD symptoms (Gaudet et al., 2021; Gaynor et al., 2009).

The early family environment may exert long-term influences on children’s neurodevelopment (Benavente-Fernández et al., 2019; Delaney et al., 2023; Hutton et al., 2015; Miller et al., 2024). For instance, we have previously shown that a cognitively stimulating environment was associated with neurocognitive development in toddlerhood and better executive function in preschool children with CHD (Bonthrone et al., 2021; Chew et al., 2024). However, little is known about the role of a stimulating home environment in shaping the behavioral outcomes of preschool children with CHD.

The aim of this study was to assess the behavioral profile of preschool children with CHD in order to test the hypothesis that preschool children with CHD have less optimal behavioral outcomes compared to control children.

## Methods

### Participants

Inclusion criteria for the CHD sample were children with critical or serious CHD who were recruited to the Congenital Heart Imaging Project (REC: 07/H0707/105) between September 2014 and January 2020 and who had surgery or intervention by cardiac catheterization within the first year of life. Critical CHD was defined as hypoplastic left heart syndrome, transposition of the great arteries (TGA), pulmonary atresia with intact ventricular septum, interruption of the aortic arch, and all infants requiring surgery within the first 28 days of life with the following conditions: coarctation of the aorta, aortic valve stenosis, pulmonary valve stenosis; tetralogy of Fallot (TOF), pulmonary atresia with ventricular septal defect, and total anomalous pulmonary venous connection. Serious CHD was defined as any cardiac lesion not defined as critical that requires cardiac catheterization or surgery before 1 year of age (Ewer et al., 2011).

Inclusion criteria for the control sample were children who participated in the Developing Human Connectome Project (dHCP; REC: 14/LO/1169) (Edwards et al., 2022)) and whose parents had consented to be approached for further research studies. We contacted parents of dHCP participants who were projected to be the same age as participants with CHD when questionnaires were completed. Exclusion criteria for both groups were children born before 31 completed weeks of gestation.

The study was approved by the National Research Ethics Committee (19/LO/0451). Parents of both children with CHD and controls were contacted when their children were between four and six years of age. Parents were emailed the study information sheet and questionnaires were posted to those who agreed to participate and provided informed written consent.

### Behavioural Assessments

Five parent-rated measures were used to evaluate children’s comprehensive behavioral profile. The Children’s Behavior Questionnaire—Very Short Form (CBQ-VSF) measured three temperamental traits: Surgency, Negative Affectivity and Effortful Control (Putnam & Rothbart, 2006). The Social Communication Questionnaire (SCQ) (Rutter et al., 2003) was used to assess autism traits; it has 40-item Yes/No questions which summate into a total score. ADHD symptoms were assessed using the ADHD Rating Scale-IV (DuPaul et al., 1998) which measured the Inattention and Hyperactivity-Impulsivity scores. The Empathy Questionnaire (EmQue) assessed the first three levels of empathy in infants’ and young children’s behaviours (Rieffe et al., 2010): Emotional Contagion, Attention to Others’ Feelings, and Prosocial Actions; it has 20 items. The Strengths and Difficulties Questionnaire (SDQ) was used to measure psychological attributes and identify potential difficulties as well as strengths (Goodman, 1997) such as emotional, conduct, hyperactivity/inattention, peer relationship problems, and prosocial behaviour. Further details of these measures are provided in the Supplementary Material (eMethods).

### Environmental Factors

The Cognitively Stimulating Parenting Scale (CSPS) was used as a covariate in our models to measure the availability and variety of experiences that promote cognitive stimulation at home and in the family. The CSPS was adapted from the Home Observation Measurement of the Environment (Bonthrone et al., 2021). We used the Index of multiple deprivation (IMD) as a measure of neighbourhood deprivation, and a proxy for parental socio-economic status. The IMD combines information from 7 domains to produce an overall relative measure of neighbourhood deprivation in England. Parents’ residential postcode at follow-up assessment was used to calculate the IMD from the 2015 data release and reported as percentile ranks. (http://imd-by-postcode.opendatacommunities.org/; Accessed December 15, 2023).

## Statistical Analysis

Data analysis was completed in R using RStudio 2023.12.0 Build 369 and SPSS version 29. Firstly, missing data (1.6%) were imputed in R using missForest (Stekhoven & Bühlmann, 2012). This method of imputation has the advantages that it has no need for tuning parameters nor does it require assumptions about distributional aspects of the data. Data were assessed for normality using density plots, QQ-plots, and Shapiro–Wilk tests using SPSS. Variables were also checked for correlation and homoscedacity. Demographic and environmental data of CHD and controls were compared using Mann-Whitney U test.

Data of all 14 behavioural variables from five behavioural measures were then corrected for age and standardised before comparing outcomes of children with CHD and controls. Outlier data were defined as any data more than 1.5 times of IQR below the first quartile, or more than 1.5 times of IQR above the third quartile (Tukey, 1977). These outliers were removed from further analysis, and the number removed from each behavioural variable is shown in Supplementary Material (eTable). In order to compare behaviour of preschool children with CHD and controls, univariate analysis of each age-adjusted behavioural variable was conducted using robust regression, controlling for sex, GA at birth, and IMD. Statistical significance was determined after false discovery rate (FDR) correction (Benjamini & Hochberg, 1995). Using the PROCESS macro for SPSS (Hayes, 2022), moderated multiple regression analyses were conducted to examine the role of group (CHD or control) in the relationship between CSPS and age-adjusted behavioural variables that were significantly different between children with CHD and controls after covarying for sex, GA and IMD.

## Results

Eighty-six parents of children with CHD agreed to receive questionnaires, and 66 were returned (77% return rate). Five children were excluded, as they were born at less than 31 weeks gestation, and 5 children did not have cardiac surgery. Data from 56 children with CHD were included in the analysis. Five children in the CHD sample had confirmed or suspected genetic abnormality (2 children had CHARGE [coloboma, heart defects, choanal atresia, growth retardation, genital abnormalities, and ear abnormalities] syndrome, 2 had 22q11 deletion, and 1 child had a suspected but not confirmed genetic abnormality).

In total, 317 parents of control children agreed to receive questionnaires and 221 were returned (70% return rate). Six children were born at less than 31 weeks of gestational age. The final control sample size was 215 children. Figure 1 shows details of participant recruitment.

**Figure 1:**
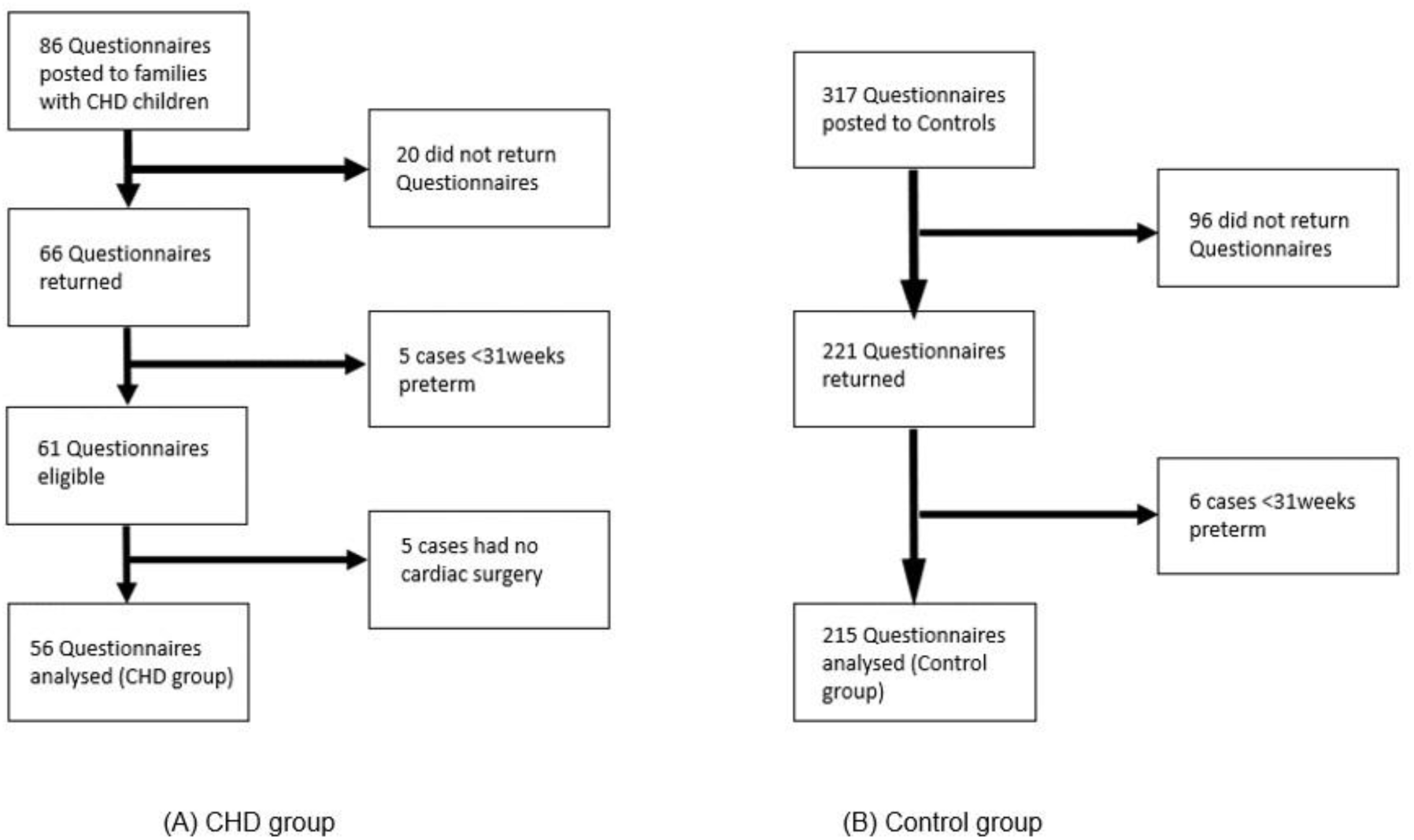
Flow chart of participant recruitment for both CHD and Control groups.

Table 1 shows participants’ demographic and environmental data, and the primary cardiac diagnoses of the children with CHD. Children with CHD were born at a younger gestational age (p < .001) and had lower CSPS scores (p = <0.001) compared to controls. Lower CSPS scores denote a less stimulating home environment. There was no difference between CHD and controls in sex distribution (p = 0.897), IMD (p = 0.756), or age at assessment (p = 0.921).

**Table 1:**
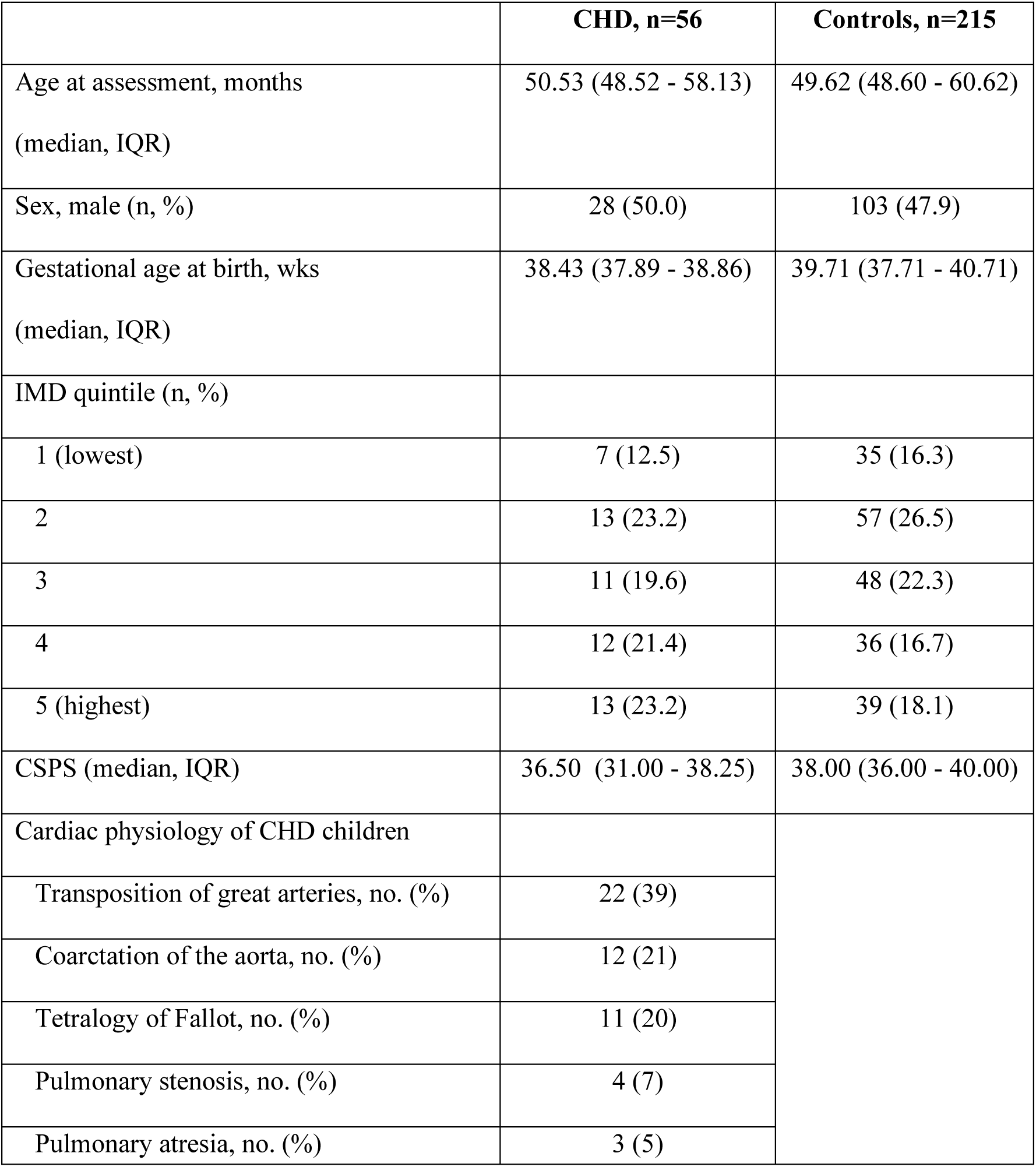
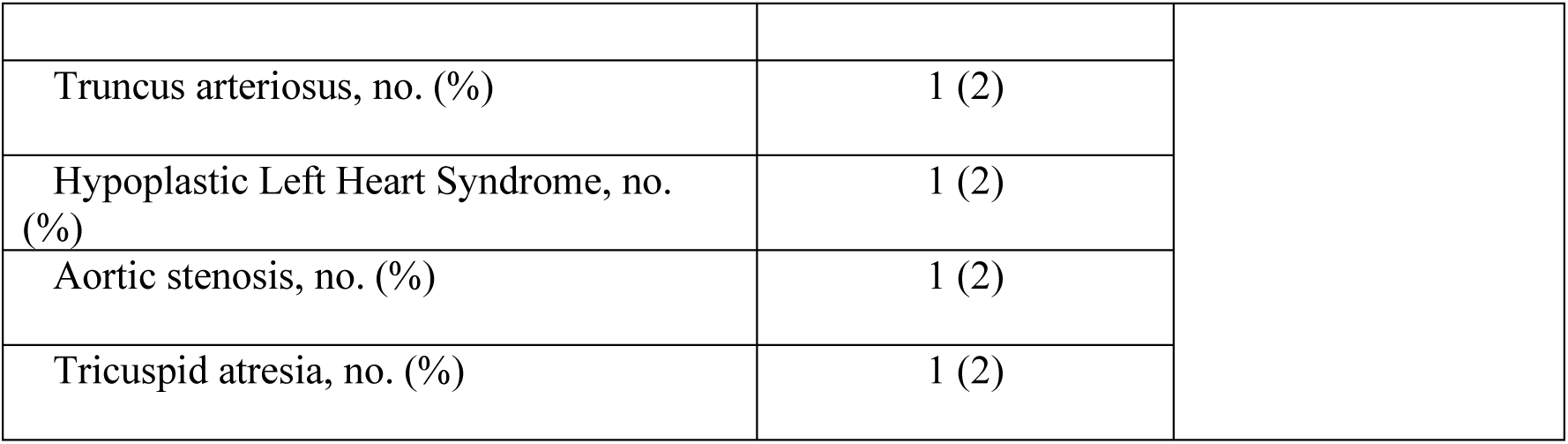
Demographic and socio-environmental data of children with CHD and controls.

Results of univariate general linear modelling (GLM) controlling for 4 covariates (group, GA at birth, Sex, IMD), showed that children with CHD had higher hyperactivity-impulsivity (ADHD-RS) (B=-0.467, p=0.023), hyperactivity/inattention scores (SDQ) (B=-0.450, p=0.023) and more peer relationship problems (SDQ) (B=-0.504, p=0.023) compared to controls. (Table 2). Lower GA at birth was associated with higher hyperactivity-impulsivity scores (B=-0.047, p=0.028), while girls had better effortful control scores (B=0.411, p=0.014).

**Table 2:** Behavioral outcomes of CHD and controls

| Outcomes<br>(n, median, IQR) | CHD | Control | $p_{FDR}$ value |
| --- | --- | --- | --- |
| Surgency (CBQ) | 55.0 (44.5 - 60.0) | 54.0 (47.0 - 60.0) | 0.762 |
| Negative Affect (CBQ) | 48.0 (40.0 - 55.0) | 46.0 (39.0 - 54.0) | 0.244 |
| Effortful Control (CBQ) | 64.0 (57.8 - 68.5) | 65.0 (60.0 - 69.0) | 0.379 |
| Inattention (ADHD-RS) | 5.0 (3.0 - 8.0) | 4.0 (1.0 - 7.0) | 0.154 |
| Hyperactivity-impulsivity (ADHD-RS) | 5.8 (4.0 - 12.0) | 5.0 (2.0 - 7.0) | 0.023* |
| Social communication (SCQ) | 5.5 (2.8 - 10.0) | 4.0 (2.0 - 7.0) | 0.093 |
| Emotion Contagion (EmQue) | 2.0 (0.0 - 4.0) | 1.0 (0.0 - 3.0) | 0.093 |
| ATOF (EmQue) | 9.0 (7.0 - 10.0) | 9.0 (8.0 - 11.0) | 0.154 |
| Prosocial Actions (EmQue) | 6.0 (5.0 - 8.0) | 6.0 (5.0 - 8.0) | 0.713 |
| Emotional problems (SDQ) | 1.0 (0.0 - 3.0) | 1.0 (0.0 - 2.0) | 0.053 |
| Conduct problems (SDQ) | 2.0 (1.0 - 3.0) | 1.0 (0.0 - 2.0) | 0.154 |
| Hyperactivity/inattention (SDQ) | 4.0 (2.3 - 6.8) | 3.0 (1.0 - 5.0) | 0.023* |
| Peer relationship problems (SDQ) | 1.0 (0.0 - 3.8) | 1.0 (0.0 - 2.0) | 0.023* |
| Prosocial behavior (SDQ) | 7.0 (6.0 - 9.0) | 8.0 (6.0 - 9.0) | 0.176 |

\* $p < 0.05$ after FDR correction, controlling for group, GA, Sex and IMD

A significant interaction between group (CHD, control) and CSPS scores was observed in predicting hyperactivity-impulsivity (ADHD) (p=0.001), hyperactivity/inattention (SDQ) (p=0.018), and peer relationship problems (SDQ) (p<0.001). CSPS scores predicted hyperactivity-impulsivity (ADHD-RS) (B=-0.092, p<0.001), hyperactivity/inattention (SDQ) (B=-0.088, p<0.001), peer relationship problems (SDQ) (B=-0.124, p<0.001) in children with CHD, but not in controls (hyperactivity-impulsivity (ADHD): B=-0.005, p=0.727; hyperactivity/inattention (SDQ): B=-0.019, p=0.225; peer relationship problems (SDQ): B=-0.002, p=0.911) (Fig 2A-C).

**Figure 2:**
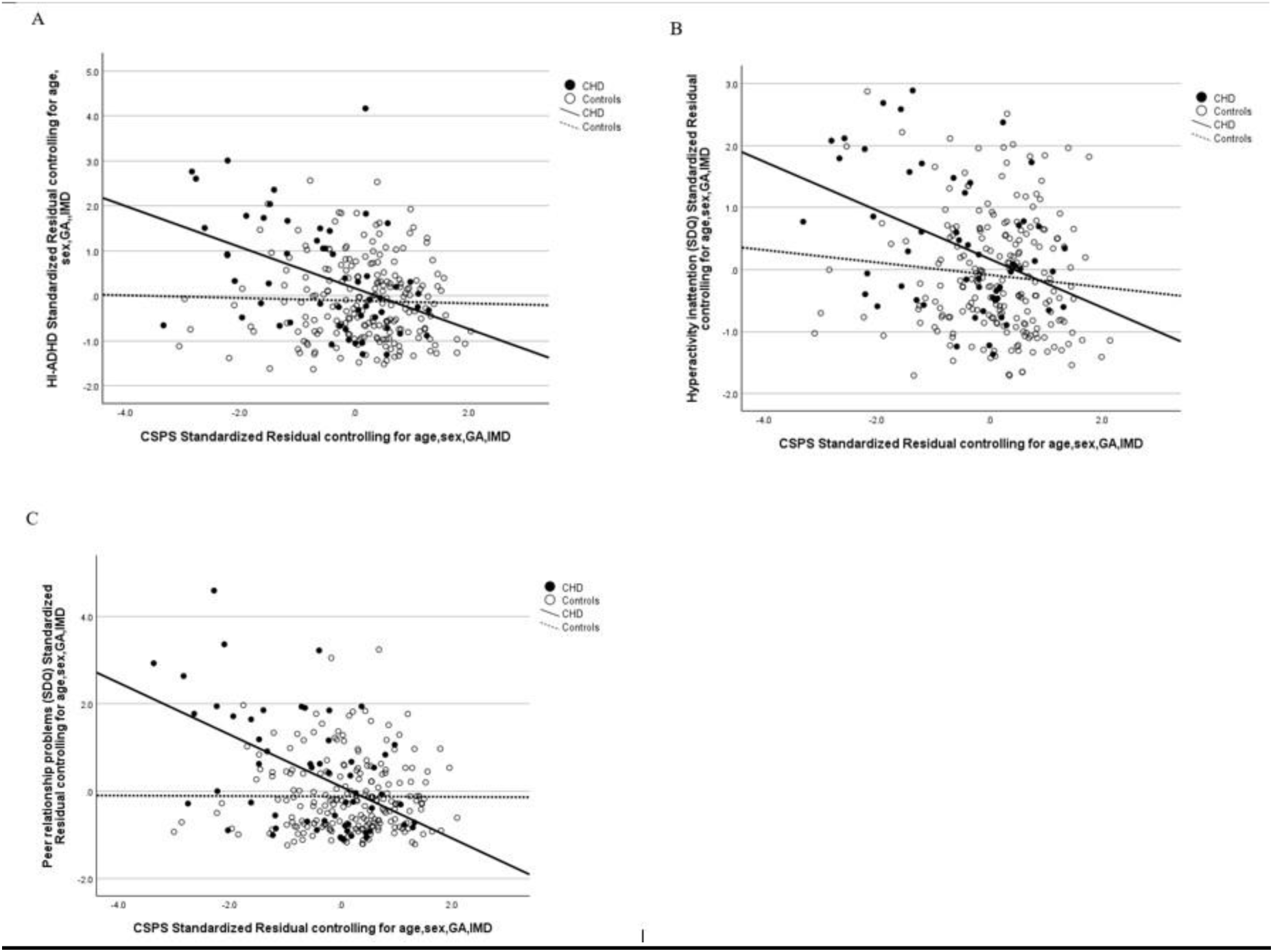
Cognitively stimulating parenting scale (CSPS) scores significantly predicted A, Hyperactivity-Impulsivity (ADHD) scores in the CHD group but not in controls, B, Hyperactivity/inattention (SDQ) scores in the CHD group but not in controls, C, Peer relationship problem (SDQ) scores in the CHD group but not in controls.

## Discussion

Our results demonstrated that preschool children with CHD exhibit more hyperactivity-impulsivity and inattention problems, and peer relationship problems than control children. We did not find any differences between children with CHD and controls in the other behavioural measures assessed. Furthermore, we found that a more cognitively stimulating home environment significantly predicted lower hyperactivity-impulsivity, inattention and peer relationship problems in preschool children with CHD but not in controls.

Studies investigating the prevalence of ADHD and the severity of ADHD symptoms in CHD have largely been conducted in school-aged children, adolescents and adults (Hasan et al., 2023). The prevalence of ADHD traits or diagnosis is higher in cohorts of children and adolescents with mixed types of CHD (Liamlahi et al., 2014; Loblein et al., 2023; Shillingford et al., 2008; Tsao et al., 2017; Werninger et al., 2020; Yamada et al., 2013) and in single physiology groups (HLHS, d-TGA, TOF) (Davidson et al., 2015; DeMaso et al., 2014; Holland et al., 2017) when compared to healthy controls or population norms. Multiple studies have shown that cyanotic CHD is a significant risk factor for ADHD diagnosis or traits (Chen et al., 2022; Hansen et al., 2012; Hövels-Gürich et al., 2007; Wang et al., 2021). However, there are only a few studies investigating ADHD traits in preschool children with CHD. A small case-control study of 12 preschool children with TGA and 30 controls did not show any between-group difference in level of hyperactivity/inattention (Hülser et al., 2007). Our results showing higher prevalence of hyperactivity, impulsivity and inattention behaviours in preschool children with CHD compared to contemporaneous controls are in line with two studies comparing ADHD traits in preschool children with CHD and population norms (Gaudet et al., 2021; Gaynor et al., 2009). Our previous work, using a comprehensive eye tracking battery to assess visual attention in toddlers (22 months) with CHD and controls, showed that toddlers with CHD had slower reaction times during selective and exogenous attention tasks (Bonthrone et al., 2024), which suggests that precursors of inattention and behavioural control difficulties could already be observed in toddlerhood.

We also found that our cohort of preschool children with CHD had more parent-rated peer relationship problems than controls. These findings are in line with previous studies that have shown young adults with atrial or ventricular septal defects have a fourfold increased risk of social interaction difficulties compared to healthy peers (Lau-Jensen et al., 2023). School-age children with CHD had previously been shown to experience peer relationship problems using the same measure we used (Werninger et al., 2020). Peer relationship problems may be partly driven by impaired capacity to recognize facial emotion expressions and identify false beliefs (Theory of Mind), which have previously been reported in school-aged children with CHD (Bellinger et al., 2011; Calderon et al., 2014; Ehrler et al., 2023).

In our study we did not find a difference in empathy or temperament in preschool children with CHD and no previous studies have measured these outcomes in similar age group. We also found no difference in autism traits between CHD preschoolers and controls when assessed using a screening tool (SCQ). This is in contrast with one other study that used SCQ (Bean Jaworski et al., 2017) and another study that used CBCL (Gaynor et al., 2009) which reported that preschool children with CHD were more likely to screen for autism and pervasive developmental disorder when compared to population norms, respectively. Further studies are therefore needed to investigate autism risk in preschool children with CHD.

This study provides further evidence of the role of a cognitively stimulating home environment in affecting outcomes of at-risk groups of children. We have previously shown that a more cognitively stimulating home environment was associated with greater cognitive scores on the Bayley Scales of Infant and Toddler Development at 22 months in toddlers with CHD (Bonthrone et al., 2021), and better executive function outcomes in our preschool cohort with CHD (Chew et al., 2024). In this study, a more cognitively stimulating home environment was associated with lower hyperactivity-impulsivity, inattention and peer relationship problems in children with CHD but not in controls, suggesting that an enriched home environment has a potentially protective and modifiable effect resulting in increased child resilience. The lack of effect in our controls suggests that children with CHD may be differentially susceptible to their environment (Belsky, 2013).

## Strengths and Limitations

A strength of this study is the inclusion of a large contemporaneous control sample. In addition, the age range of our cohort is fairly narrow within their preschool years, thus not affected by high levels of teacher-led schooling, which modulates behaviour and socioemotional development (Wang et al., 2022).

There are several limitations in this study. Our sample is from a single centre and may not be representative of the UK, or other cultures, especially where there are significant differences in how preschool children are raised (Kärtner et al., 2020). The use of parent-rated questionnaires may introduce potential common method variance bias, and future studies should compare data from parent-rated questionnaires to those obtained with objective assessments (Bennetts et al., 2016). Finally, this is a cross-sectional study, and therefore we were unable to investigate whether any significant behavioural difficulties in the preschool years are associated with future psychopathology.

## Conclusion

We have shown that preschool children with CHD have more hyperactivity-impulsivity, inattention and peer relationship problems compared to healthy controls. Our study suggests that these behavioural problems may be mitigated by a cognitively stimulating home environment. Thus environmental factors should be considered when planning intervention research aimed at improving behavioural outcomes in children with CHD.

The supplementary material for this article is in a separate document.

## Data Availability

All data produced in the present study are available upon reasonable request to the authors

