## Supplementary material for "Behavioural Outcomes of Preschool Children with Congenital Heart Disease and Controls": Supllemental File

**Supplemental Online Content**

eMethod

eTable

eReferences

### eMethod

Details of the questionnaires used are described below.

#### 1, Children's Behavior Questionnaire—Very Short Form (CBQ-VSF)

The Children's Behavior Questionnaire—Very Short Form (CBQ-VSF) is a 36-item parent-rated questionnaire assessing temperament, which is validated for children aged 3–8 (Putnam & Rothbart, 2006). The CBQ is based on Rothbart's psychobiological theory of temperament, which defined temperament as relatively stable and biologically based individual differences in levels of reactivity and self-regulation (Derryberry & Rothbart, 1988). This questionnaire asked parents to describe their children in situations occurring in everyday life using a 7-point scale from 1 (extremely untrue of your child) to 7 (extremely true of your child); thus higher scores indicate higher levels of a temperamental trait. The CBQ-VSF provides scores for three temperamental traits: Surgency, Negative Affectivity and Effortful Control. Surgency is characterised by a disposition toward positive emotions, high activity level and a rapid approach to potential rewards (Putnam et al., 2001). Negative Affectivity is characterised by shyness, discomfort, anger–frustration, fear, sadness and un-soothability. Effortful Control encompasses voluntary attentional focusing, attentional shifting, inhibitory and activational control of behaviours (Rothbart et al., 2001).

#### 2, Attention-Deficit Hyperactivity Disorder Rating Scale IV (ADHD-RS IV)

The ADHD RS-IV (DuPaul, Anastopoulos, et al., 1998) scale consists of 18 ADHD symptoms as defined in the DSM-IV, to be rated on a four-point Likert scale ranging from 0 (the symptom is "never/rarely" present) to 3 (the symptom is "very often" present). The scale is designed to generate an Inattentive Score (0–27 points on odd numbered items), a Hyperactive/ Impulsive Score (0–27 points on even numbered items), and a Total Score (0–54 points on all items) (DuPaul, Power, et al., 1998). It has been translated into many languages and remains valid for assessing the severity of ADHD symptoms in children and adolescents (Zhang et al., 2005) and longitudinally (Skogli et al., 2022), and in those with other clinical conditions (Montagna et al., 2020; Wyrwich et al., 2015) or after interventions (Dölp et al., 2020).

**3, The Social Communication Questionnaire (SCQ)**

The Social Communication Questionnaire (Rutter et al., 2003) is an autism screener which has 40-item parent-completed Yes/No questions. The SCQ was developed based on the established parental interview, the autism diagnostic interview (ADI) (Lord et al., 1994) and DSM-IV. The questionnaire can be used to evaluate anyone over age 4.0, as long as his or her mental age exceeds 2.0 years (Berument et al., 1999). The SCQ has been used in research and clinical settings, mainly in high-income settings (Chandler et al., 2007; Eaves et al., 2006; Gau et al., 2011). Recently a few studies have shown the validity of the SCQ in more rural settings in lower- and middle-income countries (Kipkemoi et al., 2025; Nwokolo et al., 2024; Ruparelia et al., 2016; Sangare et al., 2019) . As it takes approximately 10 min to answer, the SCQ is a cost-effective way to determine whether an individual should be referred for a complete diagnostic evaluation.

**4, Empathy Questionnaire (EmQue)**

EmQue is a 20-item questionnaire aimed at observing the first three levels of empathy in infants' and young children's behaviours (Rieffe et al., 2010): Emotional Contagion, Attention to Others' Feelings, and Prosocial Actions.

Hoffman had described the emotional development of children in four levels (Hoffman, 1990) and the EmQue measures the first three levels. 'Emotion Contagion' (Hatfield et al., 1993) manifests within the first year of life where infants attend to others' emotions. However witnessing someone in distress may result in a similar affective response due to automatic imitation (Decety & Jackson, 2004). Infants this young cannot yet differentiate between self and other, and act as though what happened to the other person happened to them (Vreeke & Van der Mark, 2003). 'Attention to Others' Feelings' is assumed to start at about one year of age. Infants become aware that although they feel distressed, it is not oneself but someone else who is in actual danger or pain. They become more aware of other people's emotions, and direct their attention to affective displays and concern of others. At 'Prosocial Actions' level, children become more responsive to others' emotional displays, and start to react prosocially. This ability to intervene on behalf of others during the second year of life can take a variety of forms, including helping, sharing, and comforting (Zahn-Waxler et al., 1992).

**5, SDQ**

The SDQ is a brief behavioural screening questionnaire for children and adolescents aged 2 to 17 years (Goodman, 1997). Designed to measure psychological attributes and identify potential difficulties as well as strengths, the SDQ consists of 25 items divided into five subscales: Emotional Symptoms Scale, Conduct Problems, Hyperactivity/Inattention, Peer Relationship Problems, and Prosocial Behaviour Scale. Each subscale has five items, and respondents rate each item on a three-point scale: "Not True," "Somewhat True," or "Certainly True." The scores for each subscale can be summed to provide a total difficulties score, and individual scores can help identify specific areas of concern. In community samples, multi-informant SDQs can predict the presence of a psychiatric disorder with good specificity and moderate sensitivity (Goodman, Ford, et al., 2000; Goodman, Renfrew, et al., 2000). The SDQ is freely available and being used as a research tool throughout the world in developmental, genetic, social, clinical and educational studies, and has been translated into over 80 languages. We used the Parent Version to collect data from parent's perspective.

**6, CSPS**

The Cognitively Stimulating Parenting Scale (CSPS) (Bonthrone et al., 2021; Vanes et al., 2021; Wolke et al., 2013) is a 28-item questionnaire adaptation of the Home Observation for Measurement of the Environment Inventory (Bradley & Caldwell, 1984) that is completed by parents.

The CSPS assesses the availability and variety of experiences that promote cognitive stimulation at home and in the family. This includes availability of educational toys, parental interactions such as teaching words or reading stories, and cognitively stimulating activities such as family excursions or trips (Bonthrone et al., 2021).

**eTable:** Number of outliers removed in each variable, and remaining number used during analysis

| Variables | CHD, Total n=56 | Controls, Total n=215 |
|---|---|---|
| Surgency (CBQ) | 0 removed, n=56 | 1 removed, n=214 |
| Negative Affect (CBQ) | 1 removed, n=55 | 0 removed, n=215 |
| Effortful Control (CBQ) | 2 removed, n=54 | 6 removed, n=209 |
| Inattention (ADHD-RS) | 2 removed, n=54 | 5 removed, n=210 |
| Hyperactivity-impulsivity (ADHD-RS) | 0 removed, n=56 | 6 removed, n=209 |
| Social communication (SCQ) | 2 removed, n=54 | 3 removed, n=212 |
| Emotion Contagion (EmQue) | 0 removed, n=56 | 7 removed, n=208 |
| Attention to Others' Feelings (EmQue) | 1 removed, n=55 | 5 removed, n=210 |
| Prosocial Actions (EmQue) | 1 removed, n=55 | 0 removed, n=215 |
| Emotional (SDQ) | 1 removed, n=55 | 12 removed, n=203 |
| Conduct (SDQ) | 0 removed, n=56 | 4 removed, n=211 |
| Hyperactivity/inattention (SDQ) | 0 removed, n=56 | 4 removed, n=211 |
| Peer problems (SDQ) | 0 removed, n=56 | 7 removed, n=208 |
| Prosocial behaviour (SDQ) | 1 removed, n=55 | 1 removed, n=214 |
